# Building AI Governance for a Moving Target: A Two-Year Academic Health System Experience

**DOI:** 10.64898/2026.11.26.26369100

**Authors:** Atif M. Islam, Brandon J. Dyer, Nishit Patel, Jaimie Weber, Tom Larkin, John Couris, Lillian Koslowske, Yael E. Bensoussan

## Abstract

**Objective:** We aim to describe the development, implementation, and early experience of a multidisciplinary, risk-stratification based artificial intelligence (AI) governance program within a large quaternary-care health system.

**Materials and Methods:** All AI tools submitted for governance review at Tampa General Hospital from May 2024 through June 2026 were retrospectively reviewed. The tiered governance framework is described that includes assessments of clinical and business utility, cybersecurity, ethics, compliance, risk, and educational impact. Review timelines, governance dispositions, and post-approval requirements are summarized.

**Results:** A total of 146 AI tools were submitted during the study period, with submission volume increasing substantially over time. Full Review was the most common pathway (88/146, 60.3%). Overall, 53 tools (36.3%) were approved, 54 (37.0%) were approved with stipulations, 4 (2.7%) were approved with specific pre-work requirements, and 4 (2.7%) were denied. Median time from submission to governance review and decision was 44 days (IQR, 27–88).

**Discussion:** AI governance is a dynamic process that should function beyond binary approval and rejection. Tiered review pathways and conditional approvals permit adequate oversight while enabling implementation requirements to be tailored to the potential risks intrinsic to each application.

**Conclusion:** A multidisciplinary, risk-stratified governance framework can provide a structured approach to AI oversight. As AI adoption accelerates, review and monitoring pathways will become increasingly important. The framework presented here may provide a practical model for health systems developing or refining their own AI governance programs.

## BACKGROUND AND SIGNIFICANCE

Artificial intelligence (AI) is now embedded into healthcare systems across clinical, operational, and administrative domains. Healthcare systems are rapidly incorporating the use AI tools to improve diagnostic accuracy, assist with documentation, and optimize billing among many other uses.[1] Data from the United States Office of the National Coordinator for Health Information Technology in 2024 stated that 71% of hospitals reported the integration of AI into their electronic health record, an increase from 66% in 2023.[2] Although these tools have resulted in significant workflow efficiency bonuses, the blistering adoption of these tools have outpaced the development of institutional-based oversight and monitoring programs.[3]

Standard review protocols previously employed for non-AI technology is insufficient in this modern age where AI performance depends heavily on local populations, specific workflows, implementation decisions, and user behavior.[4] Additionally, specifically with AI models is the possibility for hidden biases or hallucinations that can have unintended downstream consequences.[5] While many regulatory bodies such as the Food and Drug Administration (FDA) and National Institute of Standards and Technology (NIST) have been working to provide health systems with specific approvals, these validations do not answer whether a specific AI tool is safe, equitable, or sustainable in the institution’s local environment.

As many of these tools are commercially available, the responsibility for safely and effectively operationalizing these tools falls largely on individual health systems. Many national committees such as the World Health Organization (WHO) and Joint Commission are all increasingly recommending lifecycle oversight, risk management, local validation prior to tool implementation, and post-implementation monitoring.[6,7] The idea of AI governance was developed to directly address these concerns.

AI governance is the idea that before implementation, tools must undergo rigorous, multidisciplinary evaluation and then after approval, remain subject to post-implementation oversight to ensure safety and efficacy.[8] Governance must address all facets of the AI tool including accuracy, cybersecurity, privacy, accountability, monitoring, and the ability to implement and respond to safety signals.[9] A crucial part of this idea is that AI should be treated as a dynamic intervention rather than a static one-time software implementation.[10]

As all health systems are learning and implementing in this rapidly evolving field, reporting processes and lessons learned is of utmost importance to avoid repeating mistakes and learn from the processes others have built across the nation. Several major health systems have reported on their experiences with this challenge. University of Wisconsin reported their centralized governance process spanning from local validation to retirement whereas Duke described the regulatory best practices and lifecycle of over 50 models then later publishing their institutions “Implementation Guide”.[11–13] Massachusetts General and Memorial Sloan Kettering Cancer Center reported their institutions responsible AI-principles, assessment pathways, and post-deployment monitoring strategies.[14,15] Stanford used the Fair, Useful, and Reliable Models framework to present six AI tools and how they were adjudicated.[16] Emory reported recommendations for 13 radiology tools and the University Hospital Erlangen approved 28, rejected 9, and placed 7 AI requests on hold or under continued review.[17,18]

## OBJECTIVE

These studies establish that AI governance can be successfully operationalized but most published frameworks remain conceptual and do not include all the information required for adoption by other institutions and risk stratification guidelines are rarely reported.[19] As such, only minimal data exists to describe the process for naïve health systems to take advantage of and learn from. In this study we aim to present and report on how the AI governance framework was developed and operationalized at Tampa General Hospital, a large, quaternary, academic health system in Florida, USA. After 2 years of experience in AI governance implementation, this first paper reports on our governance framework and risk stratification methodology that has resulted in 146 tools being reviewed over a 2+ year period. This manuscript aims to report on lessons learned related to factors that shape risk stratification, how they are operationalized, and future directions and focuses to ensure safe and transparent AI deployment within our healthcare system.

## MATERIALS AND METHODS

### Study Design and AI Tool Cohort

This was a retrospective evaluation of the AI governance process at Tampa General Hospital from initiation of the program in May 2024 to June 2026. The review of this data was approved by our university’s Institutional Review Board, and the requirement to obtain informed consent was waived (USF IRB STUDY010720). All AI tools submitted for review including but not limited to clinical, research, administrative, vendor, and internally developed were included in our analysis. Each AI use case was considered individually during evaluation. Tool submissions, outcomes, and post-deployment monitoring during the study period were identified through a continuously updated institutional governance tracker.

### Health System Characteristics

Tampa General Hospital (TGH) is a private, not-for-profit academic health system that serves West Central Florida and the surrounding regions. The system is anchored by a 982-bed quaternary medical center located in Tampa, Florida and includes three community hospitals, an acute rehabilitation hospital, behavioral health teaching hospital, and an expanding network of ambulatory, emergency, surgical, and specialty care sites. Through these broad range of clinical environments, TGH is able to offer services ranging from primary care to highly specialized services including the region’s only Level I trauma center, comprehensive stroke and cancer care, solid-organ transplantation, and burn care. TGH is also the primary teaching hospital of the University of South Florida Morsani College of Medicine.

The size and heterogeneity of the health system are discussed as they directly impacted the development of the governance framework presented below. The program was developed to ensure proper review of tools for inpatient and outpatient care, diagnostic and procedural specialties, as well as operational, administrative, educational, and research tasks.

## RESULTS

### AI Governance Review Process

The AI Governance Committee (AIGC) at our institution is structured as a multidisciplinary oversight body with representation from information technology (including members with cybersecurity, analytics, informatics, and clinical engineering backgrounds), innovation, healthcare design, bioinformatics, risk management, compliance and privacy, clinical leadership, finance, ethics, and medical education. Additional as needed representation is obtained from operations (inpatient & outpatient), service line administration, or any other subject matter expert (SME) deemed to be important during the review process. This varied membership ensures thorough evaluation of our 5 principal domains: Clinical/Business Utility and Validity; Comprehensive Security Review; Ethics Review; Compliance & Risk; and Educational Impact (Figure 1). The scope of this committee includes but is not limited to tools incorporating machine learning, natural language processing, and generative AI, with the overarching goal of ensuring patient data protection, clinical efficacy, and ethical integrity.

**Figure 1.**
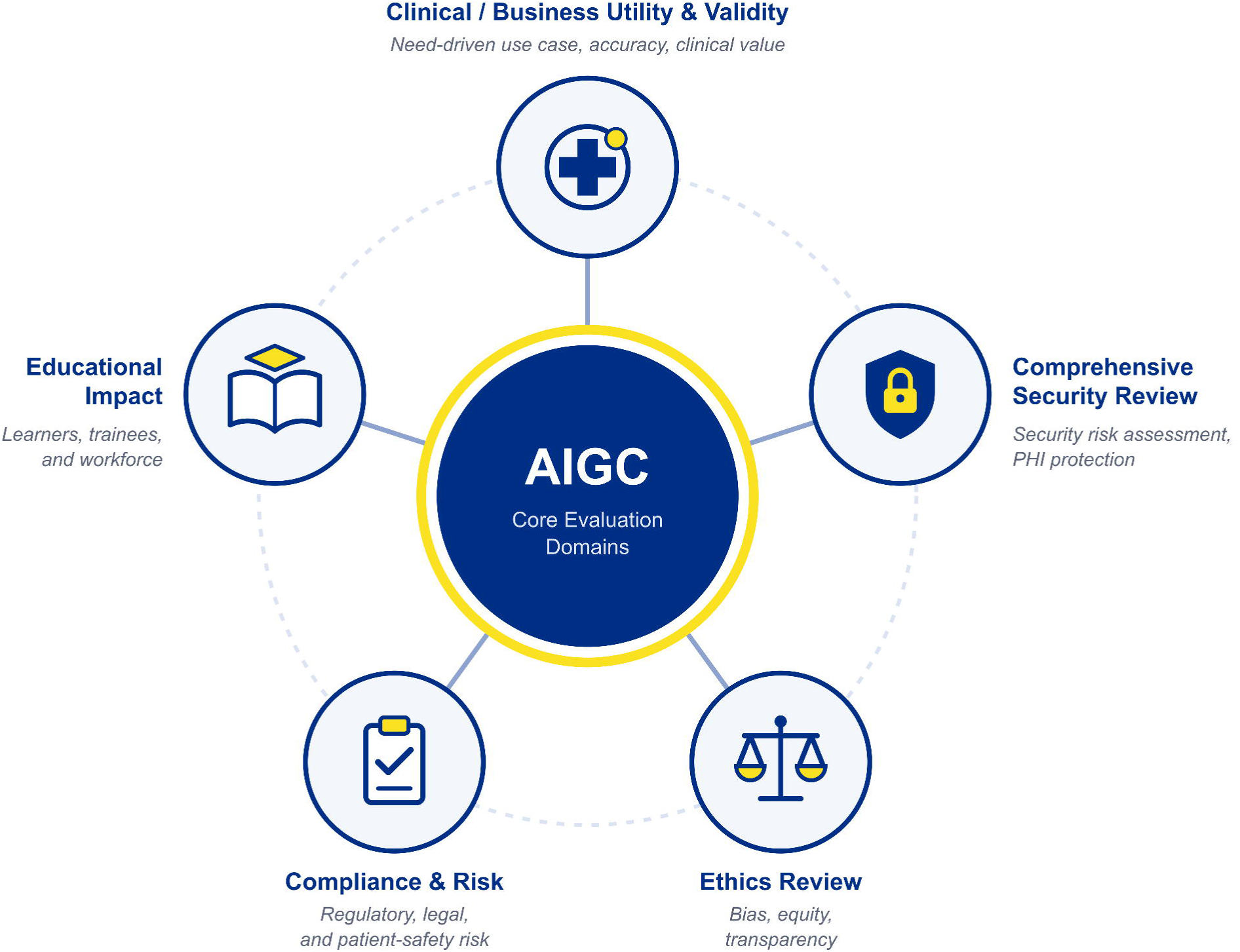
Core Domains of AI Governance Evaluation. *Alternative Text:* Diagram detailing the five core evaluation domains: clinical/business utility & validity, comprehensive security, ethics, compliance & risk, educational impact

The seven step review process begins with an individual, often a clinician or team member, who discovers a need for a new AI tool. This individual becomes the “owner” of the tool. Having a tool owner ensures that integration of the AI tool is always driven by need, clinical, operational, or otherwise. Prior to committee evaluation, the proposed tool is required to proceed through a series of pre-evaluation steps. First, the tool must be submitted for a Security Risk Assessment (SRA) where the tool is assessed by our institutional security team. For proper SRA, it is required that the tool presented has confirmed funding, a validated use case, and approval from a business sponsor; submission of the SRA is not required for Epic or Palantir based tools. Next, the tool owner or vendor must complete an AI questionnaire further detailing the purpose and use-case of the product (Supplement 1). Once these steps have been completed, a Pre-AI Governance Review Form must be submitted for SME review. The tool owner will then meet with the appropriate SME for a brief 30-minute meeting to discuss the AI components within the tool and its plan of utilization. Importantly, it is required that a vendor representative be present at this meeting to ensure technical questions are answered without delay. Tools can be placed “On Hold” at any point if steps are not completed in a timely manner.

Once a tool has an approved SRA, is it now considered AIGC Eligible for Review. At this point, each application is assigned to one of three review pathways based on anticipated risk and clinical impact: Full Review, Expedited Review, or Exempt (Figure 2). Separately, as the governance program matured, an abbreviated pathway was introduced for Epic-developed tools to streamline review of these notably high-volume applications.

**Figure 2.**
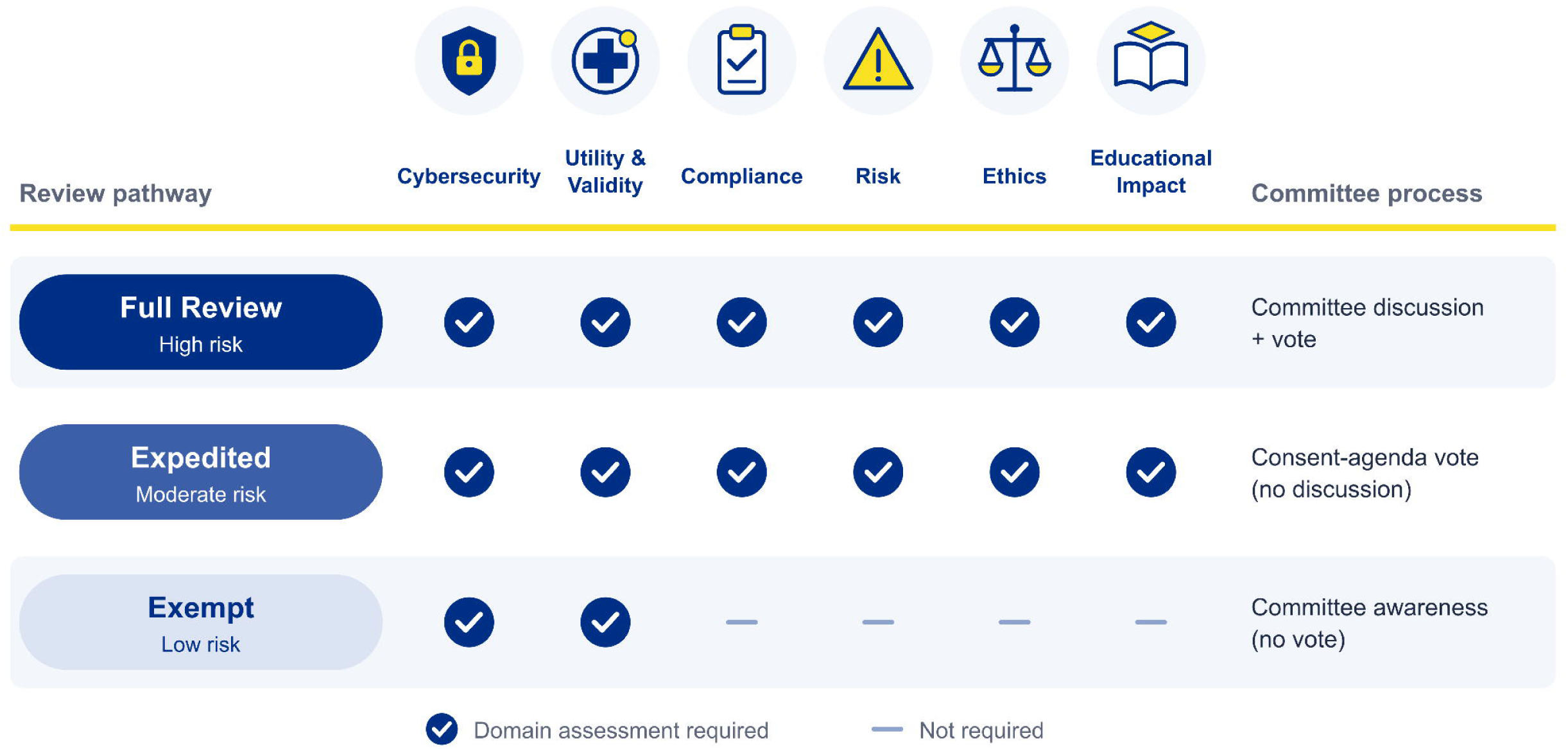
AI Governance Review Pathways and Evaluation Requirements. *Alternative Text:* Matrix comparing full, expedited, and exempt review pathways describing assessment domains and the ensuing committee process.

Full review is designed for tools with a potential for high impact/high-risk on patient privacy, outcomes, or safety, those that include PHI or could alter a patient’s treatment plan, or those that include new technologies otherwise not previously used in the healthcare setting. Full review includes complete assessments from all SME categories and a mandatory committee discussion before voting. Expedited review is for tools with moderate risk or indirect impact on patient care or operational activities. Tools under expedited review undergo assessments from all SME categories and consent-agenda voting, but do not undergo committee discussion. Exempt review is for those tools that do not require access to PHI and have no impact on patient care. Implicit to an exempt review, these tools are placed on the monthly meeting agenda for committee awareness, but do not require a vote for approval. In brief, tools that have patients as end-users, do not have a human-in-the-loop, and do not impact clinical decisions, require a full review. Our exact flowsheet is presented in Figure 3.

**Figure 3.**
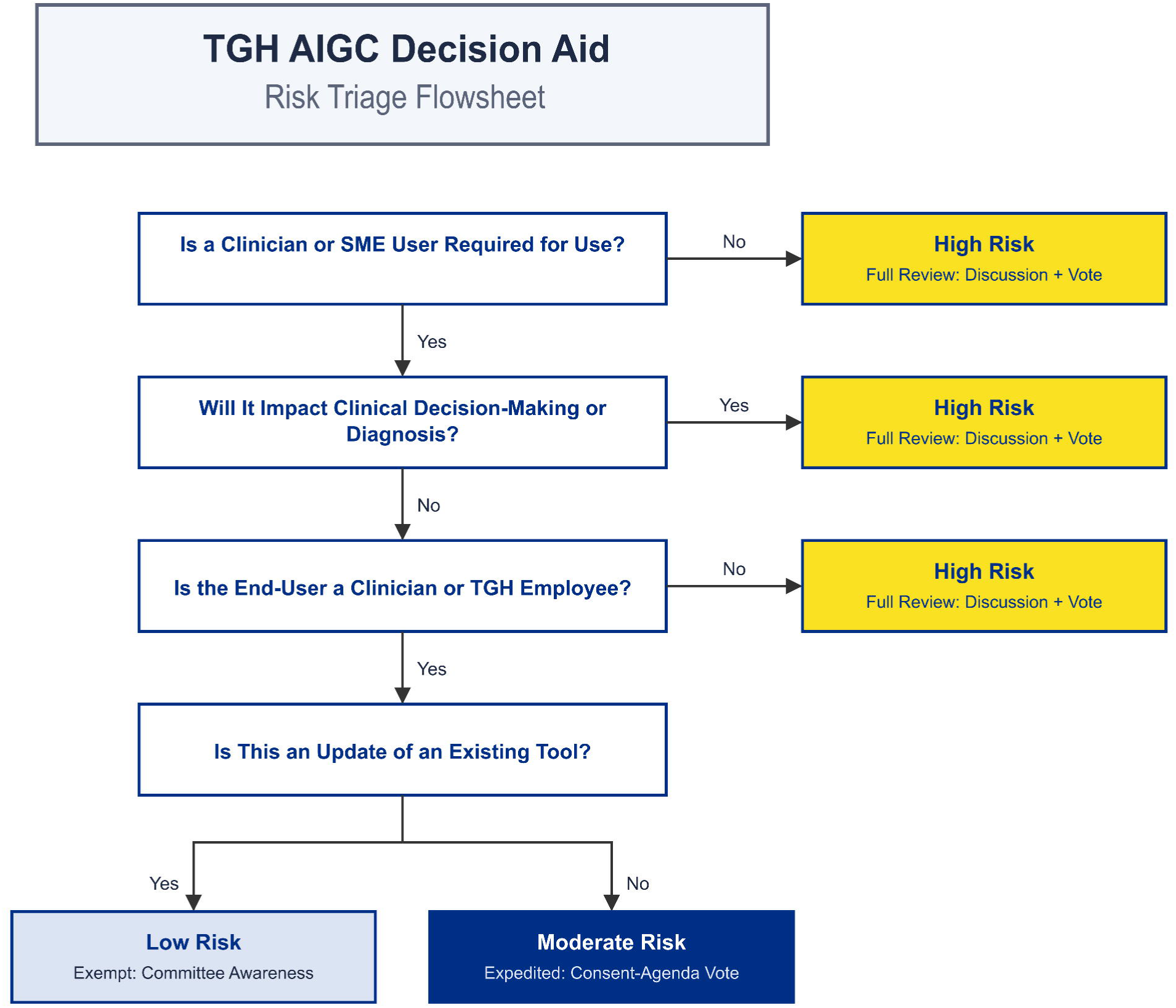
AI Tool Risk Triage Flowsheet. *Alternative Text:* Decision tree flowsheet that assigns tools to high-, moderate-, or low-risk pathways.

During the initial utility meeting, the vendor/owner meets with members of the clinical informatics and security teams to work through the flowsheet in real time and assign a preliminary review classification. This resulting classification is subject to change based on further conversations with vendors and team members. The final risk classification is solidified during an assessment team touch-base meeting that occurs prior to every monthly AIGC committee meeting. This tiered approach was intended to concentrate review time and committee resources on higher-risk tools with the greatest potential impact on patient safety and privacy. Accordingly, lower-risk tools move through the pathway substantially faster than those determined to be higher-risk. For instance, tools that are determined to be exempt receive approval from clinical informatics and proceed directly to activation.

For tools requiring governance assessment, SME’s from each of the domains individually assess the tool and provide their recommendations. Reviewers are welcome to request additional information or clarification from the tool owner or vendor during this process.

For applications requiring a committee vote, tools are placed on to the earliest available AIGC agenda. Time to presentation varies depending on if a tool becomes eligible for review in the middle of an active review cycle, the current committee backlog, and the need to prioritize time-sensitive applications or those raising leadership or security concerns.

During the monthly AIGC meeting, the completed assessments from all SME domains are presented and discussed with the tool owner present to address any outstanding questions. After discussion, voting members subsequently render a closed-door governance decision. Tools are approved based on a simple majority with a quorum of six. Decision outcomes can include approval, approval with stipulations, approval with pre-work, denial, or hold. Final decisions, including any stipulations, are communicated to the owner within one week of the meeting. Rejected tools may be resubmitted for future review if substantive changes are made that address the concerns identified during primary AIGC evaluation.

### Post-Approval Monitoring

All approved tools are subject to post-approval monitoring. Monitoring is dependent on the granted approval type.

For tools that are approved *without* stipulations, the tool owner, in collaboration with our AI Research Core Team, selects 3 specific outcome metrics that are mapped to one of our Six Outcome Pillars (Figure 4). These metrics may fall under one pillar or span multiple pillars; the goal being to ensure that each tool is evaluated on the dimensions most relevant to its function. Written 1-page reports are submitted to the AI Research Core Project Manager at 3 and 6 months post-deployment that are presented as needed at AI Tool Audit meetings.

**Figure 4.**
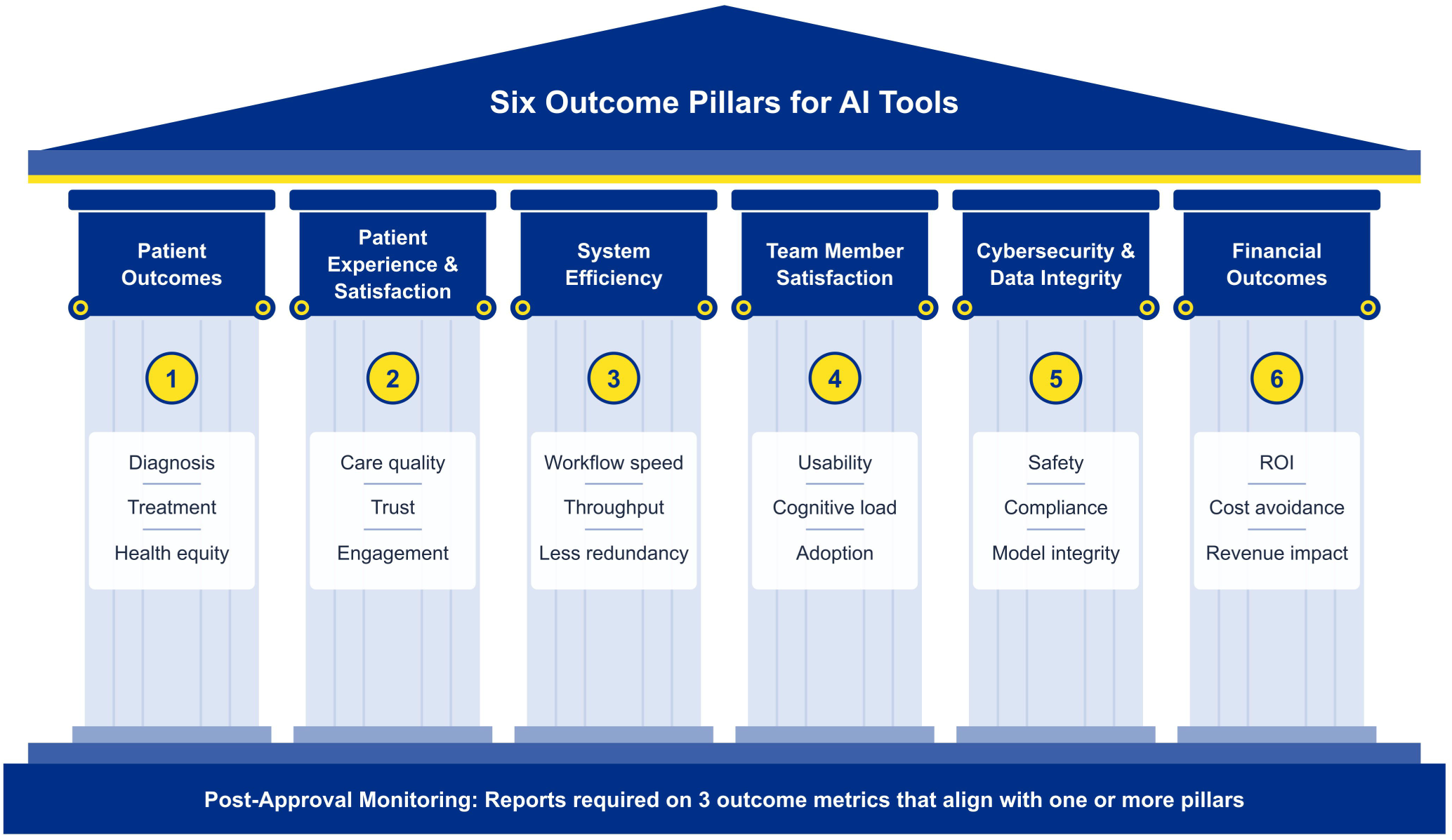
Six Outcome Pillars for Post-Implementation Monitoring. *Alternative Text:* Six-pillar framework for post-approval monitoring covering patient outcomes, patient experience & satisfaction, system efficiency, team member satisfaction, cybersecurity & data integrity, and financial outcomes.

Tools that are approved *with* stipulations must complete a structured trial before final implementation. The parameters of the trial are set by the tool owner and AIGC at the time of conditional approval; a date for re-review is also set at that time. Trials can be either time-based (3 or 6 months) or use-based (100 or 200 cases). At trial completion, a 1-page report with 3 outcomes (mapped to the aforementioned Six Outcome Pillars) and relevant accuracy metrics are submitted. At this point the tool returns to AIGC for a final decision: full approval *without* stipulations or denial.

All tool owners are required to present their monitoring reports at quarterly audit meetings convened jointly by the AIGC and the AI Research Core. In addition, at the end of each fiscal year, the AIGC and AI Research Core jointly select 3 AI tools for comprehensive in-depth analysis. Selection criteria include highest utilization volume, greatest demonstrated or anticipated impact, or a combination. These analyses provide a flagship evaluation deliverable and help guide institutional AI strategy. This longitudinal evidence base feeds directly back to the AIGC informing future approvals, modifications, and investment decisions.

### Characteristics and Disposition of AI Tools Reviewed

From May 2024 to June 2026, 146 AI tools were submitted to the AIGC for consideration. Submission volume increased substantially during the study period. The AIGC reviewed 60 submissions during all of 2025, whereas more than 60 applications were received during January-July 2026 reflecting the accelerated demand for AI tool review and use.

Median time from submission to governance review and decision during the entire study period was 44 days (IQR, 27–88 days). In August 2026, our current timelines were as follows. Mean time from submission of initial intake form to an approved SRA is 35 days. Following SRA approval, formal voting occurs within 1-2 monthly committee meetings.

Among 146 submissions, a majority were subject to full review followed by exempt, expedited, pathway not yet assigned/on hold, and denied prior to reaching the AIGC committee (Table 1). In terms of decisions, 53 (36.3%) were approved, 54 (37.0%) were approved with stipulations, and 4 (2.7%) approved with pre-work requirements. The decision to deny the tool was given in 4 (2.7%) cases, including the one denied prior to committee evaluation. Of 4 denials, two were deemed too high-risk, one lacked clinical data to support the tool’s validity, and one was not accurate enough for the use-case. The rest of the committee decisions can be viewed in Table 1.

**Table 1.** Review Pathways and Governance Decisions for Submitted AI Tools.

| **Governance Process** | **N (%)** |
|---|---|
| **Review Pathway** |  |
| Full Review | 88 (60.3%) |
| Exempt | 23 (15.8%) |
| Expedited | 22 (15.1%) |
| Pathway not yet Assigned/On Hold | 8 (5.5%) |
| Epic Tool Pathway | 4 (2.7%) |
| Denied by Security Team prior to AIGC Evaluation | 1 (0.7%) |
| **Review Decision** |  |
| Approved | 53 (36.3%) |
| Approved with Stipulation | 54 (37.0%) |
| Approved with Specific Pre-Work | 4 (2.7%) |
| On Hold | 14 (9.6%) |
| Awaiting Voting | 11 (7.5%) |
| Denied | 4 (2.7%) |
| Exempt | 3 (2.1%) |
| Preliminary Steps | 3 (2.1%) |

## DISCUSSION

### Principal Findings

This report presents a 2+ year enterprise experience with AI governance including 146 submissions. Almost all tools underwent full review, and roughly equal proportions were approved outright and approved with stipulations while very few were denied. These findings extend prior descriptions of AI governance by demonstrating how instituting a strong multidisciplinary framework early on translates into responsible-AI adoption.

We believe it important to re-state that AI governance is instituted first and foremost to protect patients and the overarching health system from *preventable* harm. In that vein, we strongly believe that governance should be a dynamic process. Our AIGC committee is not rigid in its decision tree. The frequent use of stipulations and pre-implementation requirements allows the committee to modify deployment conditions when additional safeguards are considered necessary to ensure safety.

Another important aspect of our governance process to highlight is the idea of risk-proportionate governance. We believe that not every AI application deserves the same intensity of oversight, as reflected in our separate review pathways. High-risk tools that may affect diagnosis, treatment, or any patient-related data require great scrutiny before approval. Applying that same level of scrutiny to low-risk applications would be neither efficient nor scalable in the long-term. In this sense, we are able to coexist timely innovation with patient safety.

### Scaling Governance for Next-Generation AI

As the number and complexity of AI tools entering health systems continues to increase, governance programs will need to scale their function without compromising the rigor of oversight. Reliance on fully manual review for every application is unlikely to remain sustainable making risk-based triage, tiered review pathways, and appropriate exemptions more important than ever. At our institution, the growing volume of Epic-developed tools has already prompted the development of an abbreviated AI approval pathway intended to reduce congestion issues. As the number of approved tools expand, it will also be important to develop automated systems that act as a safety net to trigger re-review of a tool if its outcomes are not aligned with institutional priorities. Finally, greater collaboration across institutions may enable development of a common taxonomy, shared model registry, and standardized vendor requirements. Together, this would enable health systems to rapidly leverage prior safety assessments and governance decisions to focus local review on whether the previously vetted tool is appropriate for the institution’s specific use-case.

### Limitations

This study should be interpreted in the light of several limitations. First, it is the experience of a single health system, and this governance framework was designed to reflect our institutional priorities; therefore, it’s structure and processes may not generalize perfectly to other organizations. Next, the framework presented here represents our current iteration of the program after multiple cycles of refinement and as such may not fully display how our review practices have changed over time. Finally, this framework has not been validated for emerging technologies such as agentic AI systems; due to their intrinsic autonomous nature, additional risk assessment may be required.

## CONCLUSION

This paper outlines a multidisciplinary, risk-stratified AI governance framework that has been instituted across a complex academic health system for over two years. It is our hope that presenting this information provides a practical starting point for other health systems seeking to establish or adapt their own AI governance program. As the volume, complexity, and autonomy of healthcare AI continues to expand, governance systems will need to become more scalable, dynamic, and collaborative. Shared standards and cross-institutional learning may be key to reducing duplicate review while preserving the rigorous oversight required for safe and effective use of AI tools.

## Data Availability

All data produced in the present work are contained in the manuscript

## ACKNOWLEDGEMENTS

CRediT Authorship Contribution Statement

We thank all study team members for their contributions to this study. A.M.I., N.P., J.W., L.K., Y.E.B. were the primary contributors to the study design. A.M.I., L.K., Y.E.B. contributed to data collection. A.M.I., B.J.D., Y.E.B. contributed to the data analysis, interpretation, and figure creation. All authors significantly contributed to the writing of the manuscript. All authors approved the final version of the manuscript and agree to be accountable for all aspects of the published work.

## Ethical Statement

The authors are accountable for all aspects of the work in ensuring that questions related to the accuracy or integrity of any part of the work are appropriately investigated and resolved. The study was conducted in accordance with the Declaration of Helsinki (as revised in 2013). Ethical approval to report this study was obtained from our university’s Institutional Review Boards (USF IRB STUDY010720).

## Declaration of Artificial Intelligence Use in Manuscript Preparation

During the preparation of this work the authors using ChatGPT 5.5 (OpenAI) to improve the readability and language of the paper. AI was not used to generate ideas, perform any statistical analysis, or create figures. After using this tool, the authors reviewed and edited the content and take full responsibility for the content of the published article.

## REFERENCES

1. Sahni NR, Carrus B. Artificial Intelligence in U.S. Health Care Delivery. N Engl J Med. Jul 27 2023;389(4):348–358. doi:10.1056/NEJMra2204673

2. Hospital Trends in the Use, Evaluation, and Governance of Predictive AI, 2023-2024 (2025).

3. Owens K, Griffen Z, Damaraju L. Managing a "responsibility vacuum" in AI monitoring and governance in healthcare: a qualitative study. BMC Health Serv Res. Sep 29 2025;25(1):1217. doi:10.1186/s12913-025-13388-z

4. Kelly CJ, Karthikesalingam A, Suleyman M, Corrado G, King D. Key challenges for delivering clinical impact with artificial intelligence. BMC Med. Oct 29 2019;17(1):195. doi:10.1186/s12916-019-1426-2

5. Subasri V, Krishnan A, Kore A, et al. Detecting and Remediating Harmful Data Shifts for the Responsible Deployment of Clinical AI Models. JAMA Netw Open. Jun 2 2025;8(6):e2513685. doi:10.1001/jamanetworkopen.2025.13685

6. Organization WH. Ethics and Governance of Artificial Intelligence for Health: WHO Guidance. World Health Organization; 2021.

7. Joint Commission and Coalition for Health AI (CHAI) Release Initial Guidance to Support Responsible AI Adoption Across U.S. Health Systems (2025).

8. Alami H, Pozelli Sabio R, Perez EJ, et al. Artificial Intelligence Governance in Health Systems: Systematic Review of Frameworks and Integrative Model Proposal. J Med Internet Res. Jun 8 2026;28:e87448. doi:10.2196/87448

9. Labkoff S, Oladimeji B, Kannry J, et al. Toward a responsible future: recommendations for AI-enabled clinical decision support. J Am Med Inform Assoc. Nov 1 2024;31(11):2730–2739. doi:10.1093/jamia/ocae209

10. Davis SE, Embi PJ, Matheny ME. Sustainable deployment of clinical prediction tools-a 360 degrees approach to model maintenance. J Am Med Inform Assoc. Apr 19 2024;31(5):1195–1198. doi:10.1093/jamia/ocae036

11. Liao F, Adelaine S, Afshar M, Patterson BW. Governance of Clinical AI applications to facilitate safe and equitable deployment in a large health system: Key elements and early successes. Front Digit Health. 2022;4:931439. doi:10.3389/fdgth.2022.931439

12. Bedoya AD, Economou-Zavlanos NJ, Goldstein BA, et al. A framework for the oversight and local deployment of safe and high-quality prediction models. J Am Med Inform Assoc. Aug 16 2022;29(9):1631–1636. doi:10.1093/jamia/ocac078

13. Economou-Zavlanos NJ, Bessias S, Cary MP, Jr., et al. Translating ethical and quality principles for the effective, safe and fair development, deployment and use of artificial intelligence technologies in healthcare. J Am Med Inform Assoc. Feb 16 2024;31(3):705–713. doi:10.1093/jamia/ocad221

14. Saenz AD, Mass General Brigham AIGC, Centi A, et al. Establishing responsible use of AI guidelines: a comprehensive case study for healthcare institutions. NPJ Digit Med. Nov 30 2024;7(1):348. doi:10.1038/s41746-024-01300-8

15. Stetson PD, Choy J, Summerville N, et al. Responsible Artificial Intelligence governance in oncology. NPJ Digit Med. Jul 4 2025;8(1):407. doi:10.1038/s41746-025-01794-w

16. Callahan A, McElfresh D, Banda JM, et al. Standing on FURM Ground: A Framework for Evaluating Fair, Useful, and Reliable AI Models in Health Care Systems. NEJM Catalyst. 2024;5(10)doi:10.1056/cat.24.0131

17. Trivedi H, Khosravi B, Gichoya J, et al. AI in Action: A Road Map From the Radiology AI Council for Effective Model Evaluation and Deployment. J Am Coll Radiol. Sep 2025;22(9):1041–1049. doi:10.1016/j.jacr.2025.05.016

18. Apfelbacher T, Koçman SE, Christoforaki M, et al. Development of an artificial intelligence (AI) governance and ethics policy: first application for the introduction of AI systems at a German University Hospital. AI and Ethics. 2026;6(3)doi:10.1007/s43681-026-01103-5

19. A. W, S. F, F. M. Governance for safe and responsible AI in healthcare organisations: a scoping review of frameworks. npj Digit Med. 2026;9(516)

