## Supplemental File 1 for "Building AI Governance for a Moving Target: A Two-Year Academic Health System Experience"

| **#** | **Question** | **Evidence Upload Requested (Y/N)** | **Answer (indicate if already provided in another questionnaire)** | **Rationale (if needed)** |
| --- | --- | --- | --- | --- |
| **Universal Questions** *The following questions establish baseline context. Additional questions apply only to certain AI classes and are shown conditionally.* | | | | |
| **Identity and Classification** | | | | |
| 1 | Provide the AI product name, current version, and release date. |  |  |  |
| 2 | Specify the AI profile. If multiple, select "Hybrid" and explain in the rationale. |  |  |  |
| 3 | For the profile(s) selected in #2, provide your Model/System Card (or equivalent) that includes the following, or answer individually: |  |  |  |
| 3a | Origin & supply chain: model origin (proprietary / open-source / third-party/foundational); how model/prompt/pipeline updates are versioned, tested, and communicated; and AI SBOM if available; otherwise provide supply-chain details (components, versions, licensing, hosting/runtime). |  |  |  |
| 3b | Purpose (≤250 words): primary business function, problem addressed, expected value, and the end-user roles/departments relying on the output. |  |  |  |
| 3c | Operational scenario: how AI is invoked; inputs; how outputs are presented; and expected user actions/decisions. |  |  |  |
| 3d | Limitations & failure modes: top limitations/warnings and how users should recognize/handle bad outputs. |  |  |  |
| 3e | Evaluation evidence: most relevant performance/quality results for this use (metrics and test conditions). For predictive models, include AUROC/F1/Sensitivity/Specificity. For generative models, provide benchmark scores (e.g., MMLU, faithfulness metrics) or human-preference evaluation results. |  |  |  |
| **Data Rights & Privacy** | | | | |
| 4 | Identify all data types that are processed, transmitted, or accessed by the AI system. If multiple, select "other" and clarify in rationale. |  |  |  |
| 5 | Would you use TGH data to train, fine-tune, or improve AI models (including foundation or third-party models)? |  |  |  |
| 6 | If yes to #2, describe the de-identification method (if applicable); if TGH data would be shared with third parties or model providers, and opt-out methods. |  |  |  |
| 7 | Define the data retention schedule for the following data types: (1) Inputs/Prompts, (2) Outputs/Responses, and (3) System Logs. Specifically, is a 'Zero Retention configuration available where no input data persists after the inference is complete? If no, justify the retention period and deletion triggers. |  |  |  |
| 8 | List all locations where data is stored, processed for inference, and backed up. Include regions, cloud providers, any cross-border data flows. Provide data flow diagram if not already provided. |  |  |  |
| 9 | Identify all subcontractors and sub-processors (including cloud hosts and model providers) that will access TGH data. |  |  |  |
| **Cybersecurity & Architecture** | | | | |
| 10 | If not already provided, include third party attestation or certification (i.e. SOC 2 Type II, ISO 27001, HITRUST). If you do not have this evidence, additional questions will be asked. |  |  |  |
| 11 | If not already provided, include evidence of a recent penetration test. Does the scope explicitly include AI-specific attack vectors (e.g., OWASP Top 10 for LLMs, Prompt Injection, Jailbreaking)? If not, provide your roadmap for adversarial testing. |  |  |  |
| 12 | If not already provided, include network architecture/data flow diagrams showing valid boundaries, external dependencies (i.e. calls to OpenAI API), and the location of any Vector Databases or Caches where 'memories' or embeddings are stored. |  |  |  |
| 13 | Indicate if Model Weight or Source Code escrow is available. If the vendor ceases operations, is there a mechanism for TGH to retrieve the trained model artifacts or the fine-tuning dataset? |  |  |  |
| 14 | Confirm that application logs and debugging tools are configured to exclude the full text of user prompts and AI outputs (to prevent PHI retention in log files), or describe the sanitization process. |  |  |  |
| 15 | Provide system-specific Business Continuity and Disaster Recovery Plan |  |  |  |
| 16 | Include AI-specific incident-response plan (addressing safety failures or hallucinations) with escalation paths. |  |  |  |
| 17 | Provide complete documentation: implementation guides, integration playbooks, validation protocols, user manuals, and release notes. |  |  |  |
| 18 | Provide breach/incident notification SLA (hours from detection). |  |  |  |
| **Clinical/Operational Safety** | | | | |
| 19 | Confirm that users are clearly informed when they are interacting with AI-generated or AI-assisted outputs; include UX copy/screenshot. |  |  |  |
| 20 | Does the system support negative feedback loops (e.g., 'Thumbs Down' or 'Report Error')? Describe how this feedback is used to retrain or correct the model. |  |  |  |
| 21 | Supply sample training materials, recommended schedule, and any CE/CME credit options. |  |  |  |
| **Ethics & Compliance** | | | | |
| 22 | Is this an FDA regulated medical device (SaMD)? If yes, provide Class and 510(k) reference number. |  |  |  |
| 23 | Describe bias detection and mitigation methods, and include Performance Parity testing or Bias-Audit Report. Does the model perform equally well across different demographic groups (e.g., age, race, gender) found in the TGH patient population? |  |  |  |
| 24 | Attach a Shared-Responsibility Matrix defining operational, security, and compliance duties among vendor, TGH, and cloud provider. |  |  |  |
| **Class A: Static, Non-Adaptive Models (Predictive ML, NLP, Computer Vision, Rules-Based only, Embedded if non-adaptive)** | | | | |
| A1 | Since the model does not learn in real-time, what specific "concept drift" triggers (e.g., changes in patient demographics, new scanner hardware) will force a mandatory model recalibration or version update? |  |  |  |
| A2 | Provide technical explainability metrics for the model's predictions (i.e. SHAP, LIME feature importance plots) to help clinical engineers validate the logic. |  |  |  |
| A3 | Confirm that the model is deterministic: will the exact same input data always result in the exact same risk score or output? If not, explain the variance. |  |  |  |
| A4 | How does the model behave when presented with input data significantly different from its training set (e.g., pediatric patient for an adult model)? Does it fail strictly or output a low-confidence score? |  |  |  |
| **Class B: Generative / Probabilistic Output Models (LLM - Conversational / Generative, LLM - RAG)** | | | | |
| B1 | Has the system undergone adversarial 'Red Teaming' to identify prompt injections, jailbreaks, or harmful content generation? If yes, attach the summary. |  |  |  |
| B2 | If TGH data is used to fine-tune a foundation model or populate a Vector Database (RAG), does TGH retain full ownership of the resulting weights and embeddings? |  |  |  |
| B3 | Does the system provide citations or 'Chain of Thought' reasoning to verify outputs? |  |  |  |
| B4 | For RAG Systems: How does the system handle queries where the answer is not found in the provided context documents? Does it explicitly state "I don't know," or does it attempt to fabricate an answer? |  |  |  |
| B5 | Does the system provide direct citations or hyperlinks to the source text (grounding) for every claim it generates? |  |  |  |
| B6 | How does the system handle patient histories that exceed the model's "Context Window"? Does it truncate the data (risk of missing recent labs) or summarize it (risk of loss of fidelity)? |  |  |  |
| B7 | What "Temperature" setting is used for clinical tasks? How do you ensure consistency in outputs (i.e. coding suggestions) when the model is inherently probabilistic? |  |  |  |
| **Class C: Decision Influencing (Decision support / recommendation engine, not autonomous)** | | | | |
| C1 | Does the AI system influence diagnosis, treatment selection, clinical prioritization, or patient-specific care decisions? If not, clarify what decisions are influenced by this system (i.e. financial, administrative). |  |  |  |
| C2 | If yes to C1, what is the IMDRF SaMD risk categorization (I, II, III, IV)? |  |  |  |
| C3 | How does the system display "low confidence" predictions? Is the user explicitly warned (i.e. color coding, icons) when the probability falls below a certain threshold? |  |  |  |
| C4 | What UX features are designed to prevent "click-through" compliance (i.e. requiring a friction step before accepting a high-risk recommendation)? |  |  |  |
| C5 | Does the system require the user to leave their primary workflow to view insights, or is it embedded? If embedded, is it passive (icon) or active (pop-up)? |  |  |  |
| C6 | How is the rationale for the recommendation displayed (i.e. SHAP plots, highlighted clinical factors, or guideline citations)? Attach a sample output. |  |  |  |
| C7 | What "Hard Rules" or safety limits constrain the model's output (i.e. "Never recommend dosage > X")? |  |  |  |
| C8 | Describe the workflow for a user to reject/override a suggestion. Does this rejection trigger a Negative Feedback Loop to update the model, or is it only logged for audit? |  |  |  |
| C9 | Describe the specific mechanisms for monitoring Harm & Accuracy Degradation (e.g., tracking reversal rates, downstream adverse events, or user complaints). |  |  |  |
| **Class D: Adaptive or Autonomous (Agentic AI, Adaptive / continuously learning systems)** | | | | |
| D1 | Suggest updating this to be: "How frequently is the model version updated by the vendor? How far in advance and in what method is the health system notified of model updates? Does the health system have the ability to control when the update is applied to allow for internal validation? |  |  |  |
| D2 | Has the system undergone adversarial 'Red Teaming' to identify prompt injections, jailbreaks, or harmful content generation? If yes, attach the summary. |  |  |  |
| D3 | Does the agent have "write" access to systems of record? If so, what hardware or logic-gated mechanisms exist to physically stop the agent if it begins executing erroneous bulk actions? |  |  |  |
| D4 | For high-impact actions, is a 'Human-in-the-Loop' confirmation strictly enforced? If an action is taken erroneously, is a 'Rollback/Undo' capability available? |  |  |  |
| D5 | Describe the mechanism for continuous learning. How is feedback validated to prevent 'Model Poisoning', and does the system require human approval before new weights are deployed? |  |  |  |
| D6 | How do you verify that new learning has not caused "Catastrophic Forgetting" or degraded performance on previously mastered baselines? Define the regression testing cadence. |  |  |  |
| D7 | List all external Tools/APIs the agent can invoke. Can TGH enforce an 'Allow/Deny' policy to restrict the agent's access to specific tools (i.e "Read-Only" access to DB)? |  |  |  |
| D8 | What safeguards exist to detect Infinite Loops or Runaway Resource Consumption? Can TGH set hard caps on API calls or Token usage to prevent financial damage? |  |  |  |
| D9 | Does the system support a forced 'State Reset' to clear the agent's memory/context if it enters a corrupted or hallucinating state? |  |  |  |
